# Janus Kinase-Inhibitor and Type I Interferon Ability to Produce Favorable Clinical Outcomes in COVID-19 Patients: A Systematic Review and Meta-Analysis

**DOI:** 10.1101/2020.08.10.20172189

**Authors:** Lucas Walz, Avi J. Cohen, Andre P. Rebaza, James Vanchieri, Martin D. Slade, Charles S. Dela Cruz, Lokesh Sharma

**Affiliations:** Department of Epidemiology of Microbial Diseases, Yale School of Public Health, New Haven, CT, 06520, USA; Section of Pulmonary and Critical Care and Sleep Medicine, Department of Medicine, Yale University School of Medicine, New Haven, CT 06520, USA; Section of Pediatric Pulmonary, Allergy, Immunology and Sleep Medicine, Dept of Pediatrics, Yale School of Medicine, New Haven, CT, 06520, USA; Department of Internal Medicine, Yale School of Medicine, New Haven, CT, 06520, USA; Department of Microbial Pathogenesis, Yale School of Medicine, New Haven, CT, 06520, USA

**Keywords:** Viral infection, Respiratory infection, Infection control

## Abstract

**Background:** Novel coronavirus (SARS-CoV-2) has infected over 17 million. Novel therapies are urgently needed. Janus-kinase (JAK) inhibitors and Type I interferons have emerged as potential antiviral candidates for COVID-19 patients for their proven efficacy against diseases with excessive cytokine release and by their ability to promote viral clearance in past coronaviruses, respectively. We conducted a systemic review and meta-analysis to evaluate role of these therapies in COVID-19 patients.

**Methods:** MEDLINE and MedRxiv were searched until July 30^th^, 2020, including studies that compared treatment outcomes of humans treated with JAK-inhibitor or Type I interferon against controls. Inclusion necessitated data with clear risk estimates or those that permitted back-calculation.

**Results:** We searched 733 studies, ultimately including four randomized and eleven non-randomized clinical trials. JAK-inhibitor recipients had significantly reduced odds of mortality (OR, 0.12; 95%CI, 0.03–0.39, p=0.0005) and ICU admission (OR, 0.05; 95%CI, 0.01–0.26, p=0.0005), and had significantly increased odds of hospital discharge (OR, 22.76; 95%CI, 10.68–48.54, p<0.00001), when compared to standard treatment group. Type I interferon recipients had significantly reduced odds of mortality (OR, 0.19; 95%CI, 0.04–0.85, p=0.03), and increased odds of discharge bordering significance (OR, 1.89; 95%CI, 1.00–3.59, p=0.05).

**Conclusions:** JAK-inhibitor treatment is significantly associated with positive clinical outcomes regarding mortality, ICU admission, and discharge. Type I interferon treatment is associated with positive clinical outcomes regarding mortality and discharge. While these data show promise, additional randomized clinical trials are needed to further elucidate the efficacy of JAK-inhibitors and Type I interferons and clinical outcomes in COVID-19.

**KEY MESSAGES:** *Key Question:* Can treatment of hospitalized COVID-19 patients with JAK-inhibitor or Type I interferon confer favorable clinical outcomes?

*Bottom Line:* Meta-analysis demonstrates that JAK-inhibitor treatment was significantly associated with favorable clinical outcomes in terms of mortality, requiring mechanical ventilation, and hospital discharge, while treatment with Type I interferon was significantly associated with decreased mortality.

*Why Read On?:* This study conducted a systematic review of human trials that treated patients with JAK-inhibitors or Type I interferon, and it elaborates on the potential benefits of administering these therapies at different moments of the disease course despite apparently opposite mechanism of action of these two interventions.

## INTRODUCTION

The spread of a highly pathogenic, novel coronavirus (SARS-CoV-2) has emerged as the deadliest pandemic since influenza in 1918 and has proved to be the ultimate challenge for public health organizations, health care providers, and governments at all levels.[1] Severe disease caused by SARS-CoV-2 (COVID-19) has strained intensive care unit (ICU) and personal protective equipment (PPE) resources around the world,[2] leading to ICU mortality rates as high as 20% in some population subsets.[3] As of July 30^th^, SARS-CoV-2 has infected over 17 million worldwide and led to the death of over 650,000.[4] Currently, only few medications have been suggested to improve the disease outcome and limit the lethal disease in susceptible populations. A small number of large-scale randomized clinical trials have been conducted so far, having demonstrated modest effectiveness for agents such remdesivir or dexamethasone.[5, 6] Additional therapeutics against COVID-19 are being explored, but there remains a lack of large scale RCTs of many potentially useful therapies, possibly missing some important therapeutics that can alter outcomes in COVID-19 patients.

Janus-kinases (JAKs) are transmembrane proteins that serve to mediate and amplify extracellular signals from growth factors and cytokines. Their inhibitors have been found to be effective in treating patients with inflammatory diseases.[7] These inhibiting drugs function by targeting specific Janus kinases. Both Baricitinib and Ruxolitinib predominantly inhibit JAK1 and JAK2.[7] JAK-inhibitors may be used to control high levels of cytokines and inflammation,[8] similar to secondary hemophagocytic lymphohistiocytosis (sHLH) caused by cytokine storm, seen in patients with severe SARS-CoV-2 infection.[9] These inhibitors have proved helpful in “off-label” indications, where excessive cytokine release plays a central role in the disease progression.[10] While the hypothesis of JAK-inhibitors successfully combating high levels of cytokine expression in SARS-CoV-2 infection has been shown in some small studies,[11] their effect on a larger population has not been investigated.

Type I interferons-α/β are proteins secreted by infected cells meant to induce antiviral states in neighboring cells and stimulate cytokine production.[12] Type I and Type III interferons have potent antiviral effects. These interferons work through activation of JAK/STAT pathway to activate a multitude of genes that are collectively known as interferon stimulated genes (ISGs). These ISGs act together to block the viral life cycle at different levels. Given the widespread expression of type I interferon receptors, they function as broad spectrum antivirals that can directly and indirectly inhibit the replication of RNA viruses at various moments in a viral life cycle through several mechanisms.[13] These interferons have been found to have positive therapeutic effects in the treatment of viral hepatitis,[14] and even past coronaviruses, such as the previous SARS and MERS outbreaks.[15, 16] Additionally, a recent investigation revealed that several severe cases of COVID-19 presented with a rare, X-chromosome loss-of-function mutation that impaired Type I interferon response,[17] while another demonstrated an association between COVID-19 severity and Type I interferon deficiency.[18] Various studies have found reasons to support the use of Type I interferons in combination with other antivirals to promote positive outcomes among patients with COVID-19, but many are restricted by the number of patients they treated with interferon.[19] Interestingly, these interferons perform their functions by activating JAK pathway.

Uncertainty and a lack of clinically proven prophylactic and therapeutic options have precipitated the periodic update of treatment guidelines for patients infected with COVID-19. As such, systematic reviews evaluating effects in larger patient populations are necessary to ascertain drug-related COVID-19 outcomes. In this meta-analysis, we evaluate Janus kinase-inhibitors and Type I interferons for their efficacy and ability to produce positive outcomes in patients infected with SARS-CoV-2.

## METHODS

This systematic review was conducted in accordance with Preferred Reporting Items for Systematic Reviews and Meta-Analyses (PRISMA).[20]

### Search Strategy and Study Quality Assessment

MEDLINE (via PubMed) and MedRxiv were searched since inception throughout July 30^th^, 2020 by three investigators (LW, AC, JV). The following terms were searched in free-text fields for JAK-inhibitors. For MEDLINE: “COVID-19” AND “JAK inhibitor” OR “Ruxolitinib” OR “Tofacitinib” OR “Fedratinib” OR “Baricitinib”. For MedRxiv: “COVID-19 JAK inhibitor” OR “COVID-19 Ruxolitinib” OR “COVID-19 Tofacitinib” OR “COVID-19 Fedratinib” OR “COVID-19 Baricitinib”. The following terms were searched in free-text fields for Type I interferons. For MEDLINE: “COVID-19”[Title] AND “interferon”[Title/Abstract] OR “IFN”[Title/Abstract]. For MedRxiv: “COVID-19 interferon” or “COVID-19 IFN”.

Three investigators (LW, AC, JV) independently screened titles and abstracts generated by the search. After selection, full electronic articles were then carefully evaluated for data extraction. Randomized studies included in the final analyses were scored by one investigator (LW) to formally assess for risk of bias utilizing the Risk of Bias (RoB) 2 tool (Supplementary Table 3).[21] Non-randomized studies included in the final analyses were scored by one investigator (LW), utilizing the Newcastle-Ottawa Scale (NOS) according to the following study characteristics: (1) representativeness of exposed cohort, (2) selection of nonexposed cohort, (3) exposure assessment, (4) outcome of interest not present at the start of the study, (5) comparability of cohorts, (6) outcome assessment, (7) adequacy of length of time before follow-up, and (8) adequacy of follow-up of cohorts (Supplementary Table 4).[22]

### Inclusion and Exclusion Criteria

We included clinical trials that utilized combination or sole JAK-inhibitor or Type I interferon (IFN-α, IFN-β) for the treatment of confirmed COVID-19 infection. For inclusion, possible studies must have compared treatment outcomes of those treated with a JAK-inhibitor or Type I interferon against a defined control group that did not receive this treatment. Selection required data with clearly indicated risk ratios or odds ratios (OR), or those that permitted their back-calculation. Inclusion necessitated that the trial be a human study accessible in English, and could include pediatric or adult studies, observational studies, retrospective cohorts, randomized clinical trials, and case reports.

Studies that utilized *in vivo* or animal studies, as well as those examining histological, pathological, and cellular mechanisms were excluded. Duplicate studies, review articles, commentaries, and proposed protocol were also excluded. Trials were excluded if they primarily examined other therapies where outcomes were unclear as to which participants received JAK-inhibitors or Type I interferons. Finally, studies were not included if they presented outcomes considered heterogenous across the review that made statistical synthesis impossible (e.g. Mean vs Median).

### Data Extraction and Data Analysis

Each full article that met inclusion criteria was carefully reviewed with the following baseline information extracted: first author, publication year, country, study type, type of JAK-inhibitor or interferon used, number of total participants, number of participants receiving JAK-inhibitor or interferon, and outcome measurements (Table 1). The outcome measurements consolidated included mortality, disease severity (mild/moderate vs severe/critical), mechanical ventilation, Intensive Care Unit (ICU) admission, discharge, and acute respiratory distress syndrome (Supplementary Table 1). Additional individual study definitions of COVID-19 disease severity are presented in Supplementary Table 2.

**Table 1.** Baseline characteristics of included studies. Included studies classifications of First Author, Year, Country, Study Type, Type of JAK-inhibitor/Type I interferon Used, Total Number of Participants, Number of Participants Receiving JAK-inhibitor/Type I interferon Used. Studies presented in alphabetical order by treatment group.

| **First Author, Year** | **Country** | **Study Type** | **JAK-inhibitor/**<br>**Interferon**<br>**Used** | **Total # of**<br>**Participants** | **N Participants**<br>**Receiving JAK-**<br>**inhibitor/Interferon\*** |
| --- | --- | --- | --- | --- | --- |
| Bronte 2020 (23) | Italy | Observational | Baricitinib | 76 | 20 |
| Cantini 2020a (24) | Italy | Retrospective Cohort | Baricitinib | 191 | 113 |
| Cantini 2020b (25) | Italy | Prospective Cohort, open-label | Baricitinib | 24 | 12 |
| Cao 2020 (26) | China | RCT | Ruxolitinib | 41 | 20 |
| Giudice 2020 (27) | Italy | RCT | Ruxolitinib | 17 | 7 |
| Chen 2020 (34) | China | Observational | IFN-alpha-2b | 291 | 132 |
| Davoudi-Monfared 2020 (28) | Iran | RCT | IFN-beta-1a | 81 | 42 |
| Du 2020 (29) | China | Retrospective Cohort | IFN-alpha | 182 | 178 |
| Estébanez 2020 (40) | Spain | Retrospective Cohort | IFN-beta-1b | 256 | 106 |
| Fan 2020 (35) | China | Retrospective Observational | IFN-alpha-1b | 53 | 19 |
| Hung 2020 (32) | China | RCT | IFN-beta-1b | 127 | 86 |
| Liu 2020 (30) | China | Retrospective Observational | IFN-alpha-2b | 10 | 9 |
| Pereda 2020 (31) | Cuba | Retrospective Cohort | IFN-alpha-2b | 814 | 761 |
| Wang 2020 (36) | China | Retrospective Cohort | IFN-alpha-2b | 446 | 242 |
| Zhou 2020 (33) | China | Retrospective Cohort | IFN † | 221 | 139 |

IFN = Interferon
\* No Study participants received JAK-inhibitor and Type I interferon
† Unclear – Used in combination with Arbidol

ORs were extracted from articles or back-calculated from the presented data. Data were analyzed using Review Manager version 5.4 (Cochrane Corporation, Oxford, United Kingdom) and the Mantel-Haenszel method. All analyzed variables are dichotomous, thus, Crude ORs, 95% Confidence Intervals (CIs) are reported. Heterogeneity was assessed using tau-squared and chi-squared tests for random effects and fixed effect models, respectively, as well as the I^2^ statistic. For I^2^ > 50%, the random effects model was used. Otherwise, the fixed effects model was utilized. An alpha of 0.05 was adopted to determine significance.

The meta-analysis results are presented on forest plots, with a study’s calculated OR plotted as a black square whose size is proportional to the weight afforded to the study. Bidirectional bars stemming from these black squares correspond to the risk estimate’s 95% CI. Diamonds were used to represent the summary OR; its center aligns with the OR and its width represents the summary 95% CI. Publication bias was assessed using funnel plots (Supplementary Figure 1; Supplementary Figure 2).

## RESULTS

The initial database search returned 731 articles. Two additional articles were added by manually searching retrieved reviews. After removing two duplicates, 698 articles were excluded following title and abstract screening by three investigators. After comprehensive evaluation of 33 full text articles, only 15 studies complied with the inclusion criteria. The majority of the studied excluded in the final step were excluded on the basis of not presenting outcome data in terms of those who did and did not receive JAK-inhibitor or interferon treatment. The remainder of excluded studied were due to a focus on JAK inhibition or interferon therapy as prophylaxis or heterogeneity in reporting of time among outcomes, precluding calculating pooled measures. Of the included studies, six were pre-prints. Overall, the 15 studies were comprised of four observational studies, six retrospective cohorts, four RCTs, and one prospective cohort. Figure 1 presents the meta-analysis flow chart and Table 1 presents the designs and characteristics of included studies.

**Figure 1.**
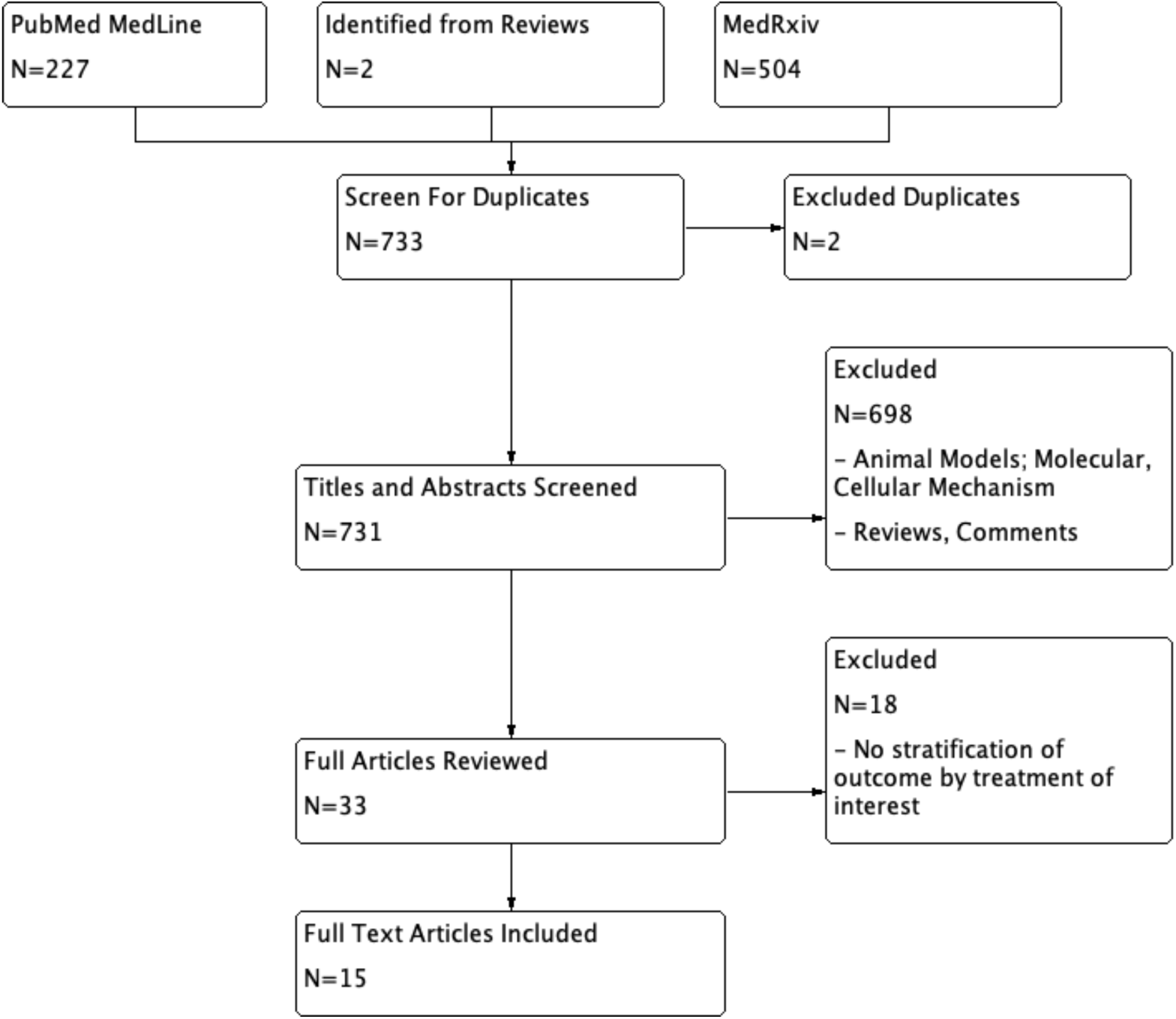
Flow diagram of study identification and assessment for eligibility. 227 and 504 studied were identified from the databases Medline and MedRxIV, respectively. Two additional articles were added by manually searching retrieved reviews. Two articles were removed as duplicates. 698 were removed after title and abstract screening not meeting inclusion criteria. 18 articles were removed after evaluation of the full article, with 15 included articles remaining.

While some studies did not report which drugs were given to which patients as standard of care, many others reported treating patients with glucocorticoids, hydroxychloroquine, chloroquine, arbidol, and lopinavir/ritonavir. All studies were conducted within a hospital setting.

### Effect of JAK Inhibition on Clinical Outcomes in COVID-19

A total of five studies investigated the effect of JAK inhibition in a controlled setting (Table 1), enrolling a total of 172 patients who received a JAK-inhibitor and 177 control participants.[23-27] The common parameters that were measured included mortality, ICU admission, requiring mechanical ventilation, acute respiratory distress syndrome (ARDS) incidence, and 14-day discharge. Meta-analysis of the five studies revealed a significantly lower odds of mortality with JAK-inhibitor (OR, 0.12; 95% CI, 0.03 – 0.39; p=0.0005), as compared to standard treatment. The effect size among the different studies demonstrated relatively little heterogeneity (I^2^=11%; Figure 2A). Pooled analyses of 2 sets of studies revealed that there was no significant association between JAK-inhibitor and COVID-19 patients requiring mechanical ventilation or developing ARDS, respectively (p>0.05; Figure 2C; Figure 2D). Both analyses included 27 patients receiving a JAK inhibitor, while the mechanical ventilation and ARDS analyses included 31 and 66 control patients, respectively. Investigation of 125 JAK-inhibitor and 90 control COVID-19 patients found that those treated with JAK-inhibitor, in comparison to those receiving standard treatment, demonstrated 0.05 (95% CI, 0.01 – 0.26) times the odds of being admitted into the ICU (p=0.0005; Figure 2B). Finally, analysis of 2 studies of 215 patients, 125 of which were treated with a JAK-inhibitor, revealed that those treated with JAK-inhibitor had significantly higher odds than those treated with standard care to be discharged at 2 weeks (OR, 22.76; 95% CI, 10.68 – 48.54; p<0.00001; Figure 2E). The analysis examining the relationship between treatment with JAK-inhibition and requiring mechanical ventilation, developing ARDS, ICU admittance, and hospital discharge demonstrated very little heterogeneity (I^2^=0).

**Figure 2.**
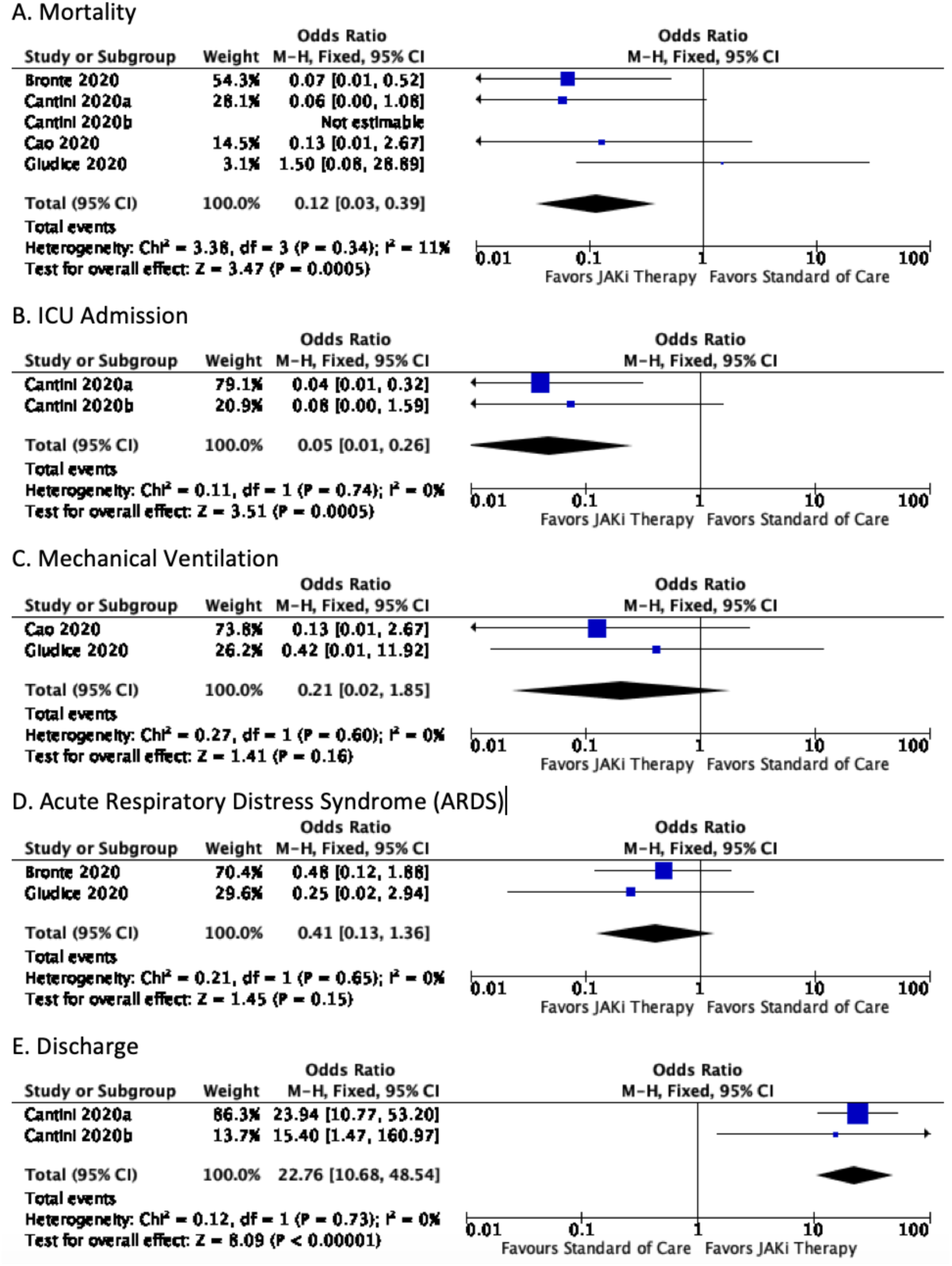
Forest plot of (A) Mortality, (B) ICU Admission, (C) Requirement of Mechanical Ventilation, (D) ARDS, and (E) Discharge of patients treated with JAK-inhibitor. The fixed effects model was used. JAK-inhibitor treatment group saw significantly reduced odds of mortality and ICU admission, as well as significantly higher odds of discharge, when compared to standard treatment. There was no significant difference between groups in regards to requiring mechanical ventilation, or the development of ARDS.

### Effect of Interferon Therapy on Clinical Outcomes in COVID-19

Meta-analysis of 3 sets of studies with 990, 454, and 1480 patients receiving Type I interferon therapy revealed that there were no significant associations between receiving Type I interferon therapy, compared to standard of care, and ICU admittance, requiring mechanical ventilation, or developing a severe or critical case of COVID-19, respectively (p>0.05; Figure 3B; Figure 3C; Figure 3D).[28-36] The analyses included 97, 167, and 537 control patients, respectively. The data exhibited very high heterogeneity in cases of ICU admittance and disease severity (both I^2^>90%), but relatively low in the case of mechanical ventilation (I^2^=12%). In the analyses of the 803 and 1415 Type I interferon receiving patients, intervention therapy was respectively associated with higher odds of being discharged (OR, 1.89; 95% CI, 1.00 – 3.59; p=0.05; N=895; Figure 3E), and significantly lower odds of mortality (OR, 0.19; 95% CI, 0.04 – 0.85); p=0.03, N=1906; Figure 3A), when compared to standard of care. The studies included in these analyses enlisted 92 and 491 control patients, respectively. Discharge data exhibited very low heterogeneity (I^2^=0%), while mortality data demonstrated very high heterogeneity (I^2^=90%).

**Figure 3.**
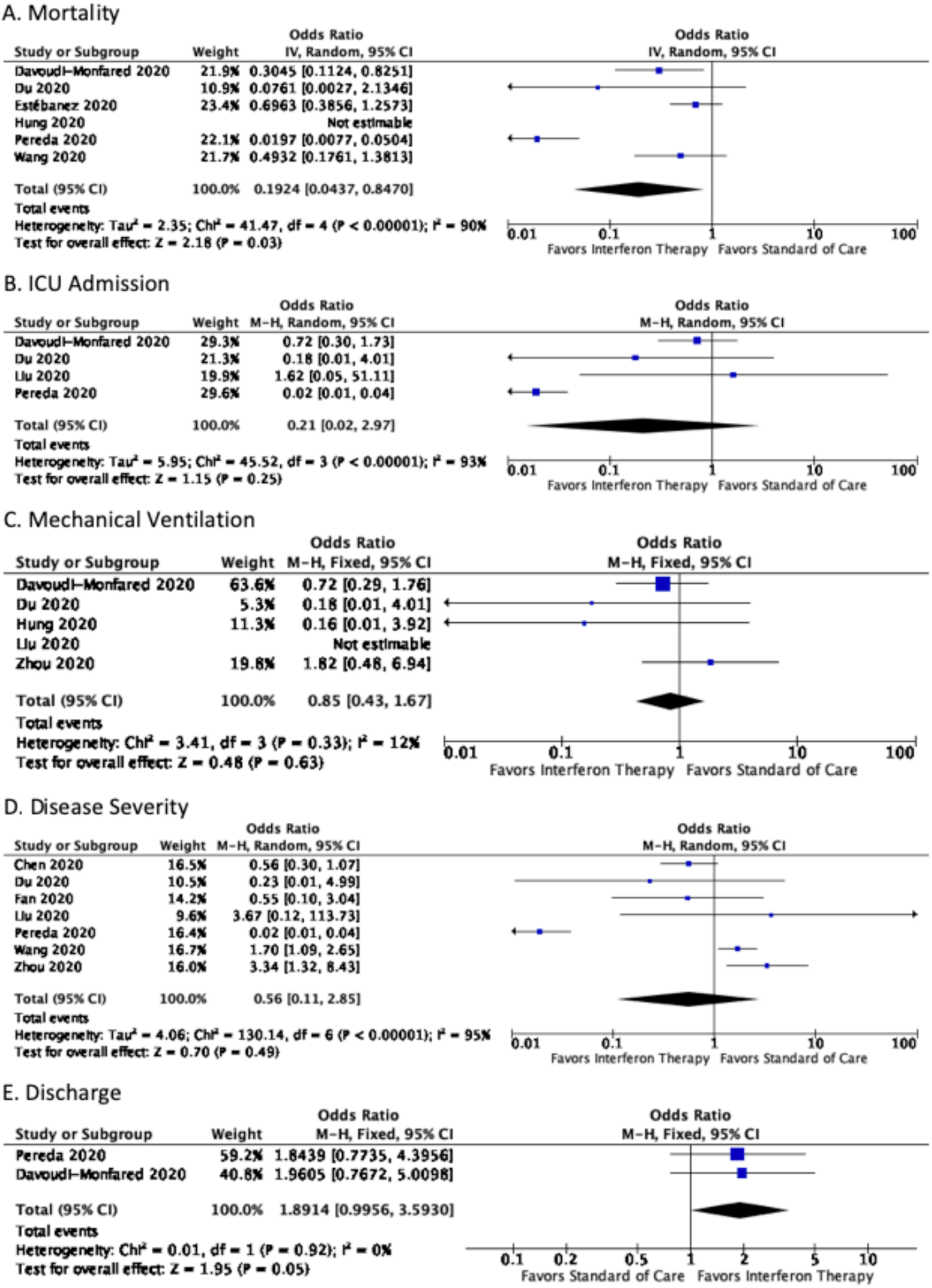
Forest plot of (A) Mortality, (B) ICU Admission, (C) Requirement of Mechanical Ventilation, (D) Severe or Critical Disease, and (E) Discharge of patients treated with Type I interferon. The fixed effects and random effects model was used dependent on the I^2^ measure of heterogeneity. Type I interferon group saw significantly reduced odds of mortality, as well as increased odds of discharge that bordered significance, when compared to standard treatment. There was no significant difference between groups in regards to requiring ICU admission, mechanical ventilation, or the development of severe or critical disease.

## DISCUSSION

As SARS-CoV-2 continues to infect millions and kill thousands daily, there is an urgent need to find novel therapies that can effectively limit COVID-19 severity. Type I interferon therapy as well JAK inhibitors represent paradoxical approaches to treat COVID-19. While Type I interferon therapy aims to limit the viral replication at the early time points to limit the subsequent disease, JAK-inhibitors aim to limit the overt inflammation that may be detrimental to the host and cause systemic inflammatory response.

However, no major randomized clinical trials have been performed to determine their efficacy in limiting the disease severity in COVID-19.

To our knowledge, this is the first systematic review and meta-analysis to investigate the role of Janus kinase-inhibitor or Type I interferon on clinical outcomes in patients with COVID-19. The results suggest a robust association between JAK-inhibitor and significantly decreased odds of mortality and ICU admission, as well as significantly increased odds for 14-day patient discharge. Furthermore, a significant association between Type I interferon and reduced mortality was also found, in addition to an association with hospital discharge bordering significance. These results suggest the potential benefit of these therapeutic options for COVID-19.

Although this study presents evidence of JAK-inhibitors and Type I interferon therapies for COVID-19 patients, the evaluated studies included conflicting results; Giudice et al. reported a positive association between JAK-inhibitor therapy and the odds of mortality,[27] while the other studies analyzed and the summary statistic calculated demonstrated a negative association between JAK-inhibitor intervention and mortality (23, 24, 26). In addition, two studies consistently demonstrated opposite associations between Type I interferon therapy and clinical outcomes,[30, 33] when comparing summary statistics and other included studies.[28, 29, 31, 34, 35] Heterogeneity among populations studied may play a role in the disparate individual results, as half of these studies were conducted in China, one was conducted in Iran, five were conducted in Western Europe, and one was conducted in Cuba. Other irreconcilable factors that may have influenced patient outcomes included individual study exclusion criterion, as well as the dosage and delivery method of the intervention.

Furthermore, as recent findings have shown that persistent viral presence contributes to disease severity,[37] the timing of the administration of both interventions may be of utmost importance. As JAK inhibitors attenuate JAK signaling and subsequent cytokine release, their administration may best be suited for patients with progressing COVID-19 who have not yet experienced a cytokine storm.[38] By contrast, as Type I interferons induce cellular antiviral states via the JAK/STAT pathway, its administration may be most efficacious early on in disease progression where the virus is still replicating. While the literature surrounding this is sparse, one study included in this meta-analysis concluded that early administration of interferon-alpha-2b could induce positive outcomes in COVID-19 patients compared to standard treatment, while its late administration was associated with slower recovery.[36]

It is important to highlight that this meta-analysis attempted to overcome the challenges posed by studies with insufficient power to detect an effect between JAK-inhibitor or Type I interferon treatment and clinical outcomes, as half of the included studies in this analysis utilized sample sizes less than 100.[23, 25-28, 30, 35] Nevertheless, despite the broad range of sample sizes and populations, the screening step of our analysis predominantly resulted in low effect size heterogeneity as evidenced by the I^2^ statistics displayed in Figure 2 and Figure 3.

This study contained no restrictions regarding study type in the exclusion criteria and, as such, many of the studies included are of retrospective design. Accordingly, baseline characteristics of patients cannot be ignored, especially as factors such as age, gender, and pre-existing comorbidities have been found in meta-analyses to be linked to negative clinical outcomes, including mortality, among COVID-19 patients.[39] One study in particular contained a large disparity in the distribution of chronic conditions across those who received Type I interferon therapy and controls.[31] In addition, these non-randomized studies are inherently limited in their ability to deduce the causality of association between the treatments of interest and clinical outcomes; they should be interpreted with caution. Another limitation of this study is the aspect of drug combination. Included studies varied in the drugs administered in the control arm and in addition to JAK-inhibitor or Type I interferon in the treatment group. Types and doses of JAK-inhibitors and Type I interferons also differed across studies. Moreover, publication bias may have been present in some of the analyses conducted (Supplemental Figure 1; Supplemental Figure 2). The low number of studies make it difficult to assess asymmetry in funnel plot analyses. However, we attempted to mitigate this bias with the inclusion of six unpublished articles. A further limitation of this study is the exclusion of a large number of studies that presented heterogenous data that precluded pooled analyses. Lastly, this meta-analysis included two studies consisting of similar study teams that examined the same association,[24, 25] enhancing the likelihood of bias in the same direction in analyses where both of these studies were included.

In conclusion, this meta-analysis supports the value of JAK-inhibitor and Type I interferon therapy as antivirals in combating SARS-CoV-2 infection. This study consolidates existing data and reaffirms the conclusion that, within COVID-19 patients, JAK-inhibitor treatment is significantly associated with positive clinical outcomes in terms of mortality, ICU admission, and discharge, as well as Type I interferon treatment’s association with positive clinical outcomes in regard to mortality and discharge. Although these findings should assist physicians deciding which antivirals to administer to SARS-CoV-2 infected patients, they also point to a clear need of additional well-designed RCTs examining the relationship of JAK-inhibitor and Type I interferon and clinical outcomes of COVID-19 patients.

## Data Availability

All data referred to in the manuscript are available online on webpages, journals, or MedRxiv.

## FUNDING

Lokesh Sharma is supported by Parker B Francis Fellowship. Charles Dela Cruz is supported by Veterans affairs Merit Grant (BX004661), Department of Defense grant (PR181442), and a U19 supplement for this work (AI089992-09S2).

## CONTRIBUTORSHIP STATEMENT

Lucas Walz (LW); conception of investigation, planning of investigation, data retrieval, article screening, data analysis, written reporting, data interpretation.

Avi J. Cohen (AC); planning of investigation, data retrieval, article screening, data analysis, written reporting, data interpretation.

Andre P. Rebaza, MD (AR); written reporting, data interpretation.

James Vanchieri (JV): data retrieval, article screening.

Martin D. Slade, MPH (MS): data analysis.

Charles S. Dela Cruz, MD, PhD (CDC): planning of investigation, data retrieval, written reporting, data interpretation; supervision.

Lokesh Sharma, PhD (LS): conception of investigation, planning of investigation, written reporting, data interpretation; supervision.

## Notes

### Competing Interest Statement

The authors have declared no competing interest.

### Author Declarations

The study conducted was a meta-analysis and thus did not require IRB approval.

